# Peroral Endoscopic Myotomy Versus Laparoscopic Heller Myotomy for Achalasia: A Meta-Analysis of Randomized Controlled Trials

**DOI:** 10.1101/2025.09.16.25335767

**Authors:** Harshawardhan Dhanraj Ramteke, Sanapala Keerthi, Ashesh Das, Bodanam Shreya, Shameek Paul, Bharadwaj Jilakaraju, Bodipudi Vineetha, Virushnee Senthilkumar, Ambala Madhulika, Syeda Hafsa Noor-Ain, Rakhshanda Khan

## Abstract

**Introduction:** Achalasia is a rare primary esophageal motility disorder characterized by impaired lower esophageal sphincter relaxation and dysphagia. Laparoscopic Heller myotomy (LHM) has long been the standard treatment, while peroral endoscopic myotomy (POEM) has emerged as a minimally invasive alternative. Comparative evidence from randomized controlled trials (RCTs) remains limited, and outcomes such as gastroesophageal reflux disease (GERD) and clinical remission require clarification.

**Methods:** We systematically searched PubMed, Embase, Cochrane CENTRAL, and Web of Science to September 2025 for RCTs comparing POEM and LHM in adult patients with achalasia. Data on demographics, previous treatment, dysphagia improvement, GERD incidence, clinical remission, and mortality were extracted. Pooled odds ratios (OR) with 95% confidence intervals (CI) were calculated using a random-effects model in Stata 18.

**Results:** Seven RCTs involving **900 patients** (465 POEM; 465 LHM) were included. Dysphagia improvement was similar between groups (log OR 0.14; 95% CI −0.32 to 0.59; *p* = 0.55). GERD incidence was higher after POEM but not statistically significant (log OR 0.59; 95% CI −0.08 to 1.25; *p* = 0.08). Clinical remission showed a non-significant trend favoring POEM (log OR 0.39; 95% CI −0.06 to 0.84; *p* = 0.09). Reduction in pH levels significantly favored LHM (log OR 0.75; 95% CI 0.18 to 1.33; *p* = 0.01). No mortality was reported.

**Conclusion:** POEM and LHM provide comparable dysphagia relief and clinical remission in achalasia. However, POEM is associated with higher GERD risk, particularly on pH monitoring. Treatment choice should balance efficacy against reflux risk, with careful long-term follow-up.

## Introduction

Achalasia is a rare but disabling esophageal motility disorder characterized by failure of the lower esophageal sphincter (LES) to relax and loss or discoordination of esophageal peristalsis. Clinically, patients suffer from progressive dysphagia to both solids and liquids, regurgitation, chest pain, weight loss, and in severe cases, pulmonary complications or mega□esophagus if untreated. The global epidemiology has only recently become clearer: pooled estimates indicate an incidence of ∼ 0.78 cases per 100,000 person□years (95% CI 0.64–0.93) and a prevalence of ∼ 10.8 per 100,000 (95% CI 8.15–13.48) [1]. Although these numbers remain low compared to many chronic disorders, achalasia imposes substantial burdens—on quality of life, nutritional status, healthcare resource use, and long□term risk of complications [2]. Over decades, the standard of care for achalasia has evolved. Laparoscopic Heller myotomy (LHM), often combined with a partial fundoplication, has been the benchmark surgical treatment, with many centers reporting durable symptomatic relief, acceptable morbidity, and low mortality. Conversely, more recently developed, peroral endoscopic myotomy (POEM) offers a minimally invasive alternative: through a submucosal tunnel, the circular (and sometimes longitudinal) muscle fibers of the esophagus and LES are divided endoscopically. POEM avoids external incisions, may allow for longer esophageal myotomy tailored to the achalasia subtype (e.g., type II vs type III), and is associated with reduced pain and quicker recovery in several reports [3,4,5].

Both LHM and POEM have shown high rates of clinical success in terms of dysphagia relief, with many studies reporting improvement in Eckardt scores, decreased LES pressures, and low rates of immediate complications. A landmark randomized controlled trial (Werner et al., 2019) showed POEM to be non□inferior to LHM + Dor fundoplication at 2-year follow-up in controlling symptoms [6]. In addition, recent long-term outcome data suggest that POEM maintains clinical success rates of around 80-85% over 3-5 years [7,8]. Nevertheless, while several systematic reviews and meta-analyses have attempted to compare POEM and LHM, there remain important limitations. The meta-analysis by Schlottmann et al. (2017) found that POEM was slightly more effective in dysphagia relief in the short term but was associated with significantly greater risk of gastroesophageal reflux disease (GERD), both symptomatic and objective (via esophagogastroduodenoscopy [EGD] and pH monitoring). However, this and other meta-analyses are hampered by heterogeneity in patient populations (treatment-naïve vs previously treated), variations in follow-up duration (POEM studies generally had much shorter follow-ups than LHM studies), inconsistent definitions of outcomes (especially for GERD), and predominance of nonrandomized observational designs [9,10]. For example, in Schlottmann et al., follow-up for POEM averaged ∼16.2 months versus ∼41.5 months for LHM, raising the possibility that longer□term complications (e.g. reflux, treatment failures) are under-ascertained [11].

Moreover, many studies do not sufficiently stratify by achalasia subtype (types I, II, III), do not control for learning curve effects (POEM is a newer method), and often do not include standardized or objective assessments of GERD (e.g. pH monitoring or histologic esophagitis). Cost, postoperative quality of life, reintervention (i.e. the need for redo myotomy or dilation), and long-term adverse effects such as Barrett’s esophagus are underreported. These limitations reduce the confidence with which clinicians can judge long-term trade-offs between efficacy and potential harms when choosing POEM vs LHM.

In light of these gaps, there is a pressing need to focus on randomized controlled trials and high-quality comparative studies with longer follow-up, consistent outcome definitions, and detailed reporting of adverse events. Such data will better clarify whether POEM truly matches or surpasses LHM in both benefits and risks, especially in domains such as reflux, reinterventions, and durability of symptom control.

Therefore, in this meta-analysis based on core randomized controlled trials (and high□quality RCT subsets), we aim to compare POEM versus LHM for adult achalasia in terms of (i) symptomatic relief (e.g., dysphagia / Eckardt score), (ii) safety and adverse events, including GERD (both symptomatic and objective), (iii) reintervention rates, and (iv) long-term durability (minimum follow-up 2-5 years). We hypothesize that while POEM will be non-inferior in symptom relief, it may carry higher rates of pathologic reflux; and that longer-term RCT□driven data will help delineate whether those trade-offs are acceptable or whether surgical myotomy with fundoplication remains superior in specific subgroups.

## Methods

### Literature Search

A comprehensive literature search was conducted in PubMed, Embase, Cochrane CENTRAL and Scopus from inception to September 2025. Search terms included “achalasia,” “peroral endoscopic myotomy,” “POEM,” “laparoscopic Heller myotomy,” “LHM,” and “randomized controlled trial.” Boolean operators and MeSH terms were applied where appropriate. References of relevant articles and previous systematic reviews were hand-searched to ensure completeness. Only randomized controlled trials directly comparing POEM with LHM were included. No language restrictions were applied, and conference abstracts, case series, and noncomparative studies were excluded. It followed PRISMA guidelines and also registered with Prospero with number CRD420251148413 [12].

#### Study Selection and Data Extraction

Two reviewers independently screened titles, abstracts, and full texts of all identified records, with disagreements resolved through consensus or third-party adjudication. Eligible studies were randomized controlled trials directly comparing POEM with LHM in adult patients diagnosed with achalasia. Noncomparative designs, case reports, and reviews were excluded.

Data were extracted in duplicate using a predesigned template. Extracted variables included study identifiers (author, year, country, sample size), baseline demographics, history of prior treatments for achalasia, and primary outcomes of interest: improvement in dysphagia, incidence of gastroesophageal reflux disease (GERD), and all-cause mortality during follow-up.

### Statistical Analysis

All statistical analyses were conducted using Stata version 18 (StataCorp LLC, College Station, TX, USA). Dichotomous outcomes were summarized as odds ratios (ORs) with 95% confidence intervals (CIs). A random-effects model (DerSimonian–Laird method) was applied to account for between-study variability. Heterogeneity was quantified using the I^2^ statistic, with thresholds of 25%, 50%, and 75% representing low, moderate, and high heterogeneity, respectively. Potential publication bias was explored through funnel plot inspection and Egger’s regression test where sufficient studies were available. A two-tailed *p* value <0.05 was considered statistically significant.

#### Risk of Bias Assessment

The methodological quality of included randomized controlled trials was assessed independently by two reviewers using the Cochrane Risk of Bias 2.0 (RoB 2.0) tool [13]. Domains evaluated included randomization process, deviations from intended interventions, missing outcome data, measurement of outcomes, and selective reporting. Each domain was rated as “low risk,” “some concerns,” or “high risk” of bias, and an overall judgment was assigned to each study. Disagreements were resolved by consensus or discussion with a third reviewer.

### Certainty of Evidence (GRADE)

The certainty of evidence for each outcome—previous treatment history, dysphagia improvement, GERD incidence, and mortality—was assessed using the GRADE (Grading of Recommendations Assessment, Development and Evaluation) framework. Evidence quality was rated across five domains: risk of bias, inconsistency, indirectness, imprecision, and publication bias. The certainty of evidence was classified as high, moderate, low, or very low. Summary of Findings (SoF) tables were generated to present pooled effect estimates with corresponding GRADE ratings for each primary outcome.

## Results

### Demographics

The initial search across PubMed, Embase, Cochrane CENTRAL, and Web of Science yielded 532 records. After removal of duplicates and screening of titles and abstracts, 48 studies were retrieved for full-text review.

Of these, 7 randomized controlled trials met the inclusion criteria and were included in the final quantitative synthesis Figure 1. The seven included trials enrolled a total of 900 patients with achalasia, of whom 510 were male and 420 were female. The pooled population was evenly distributed between the two intervention arms, with 465 patients undergoing peroral endoscopic myotomy (POEM) and 465 undergoing laparoscopic Heller myotomy (LHM). The mean follow-up across studies ranged from 12 to 60 months, with most trials reporting structured clinical and endoscopic assessments. Table S1.

**Figure 1.**
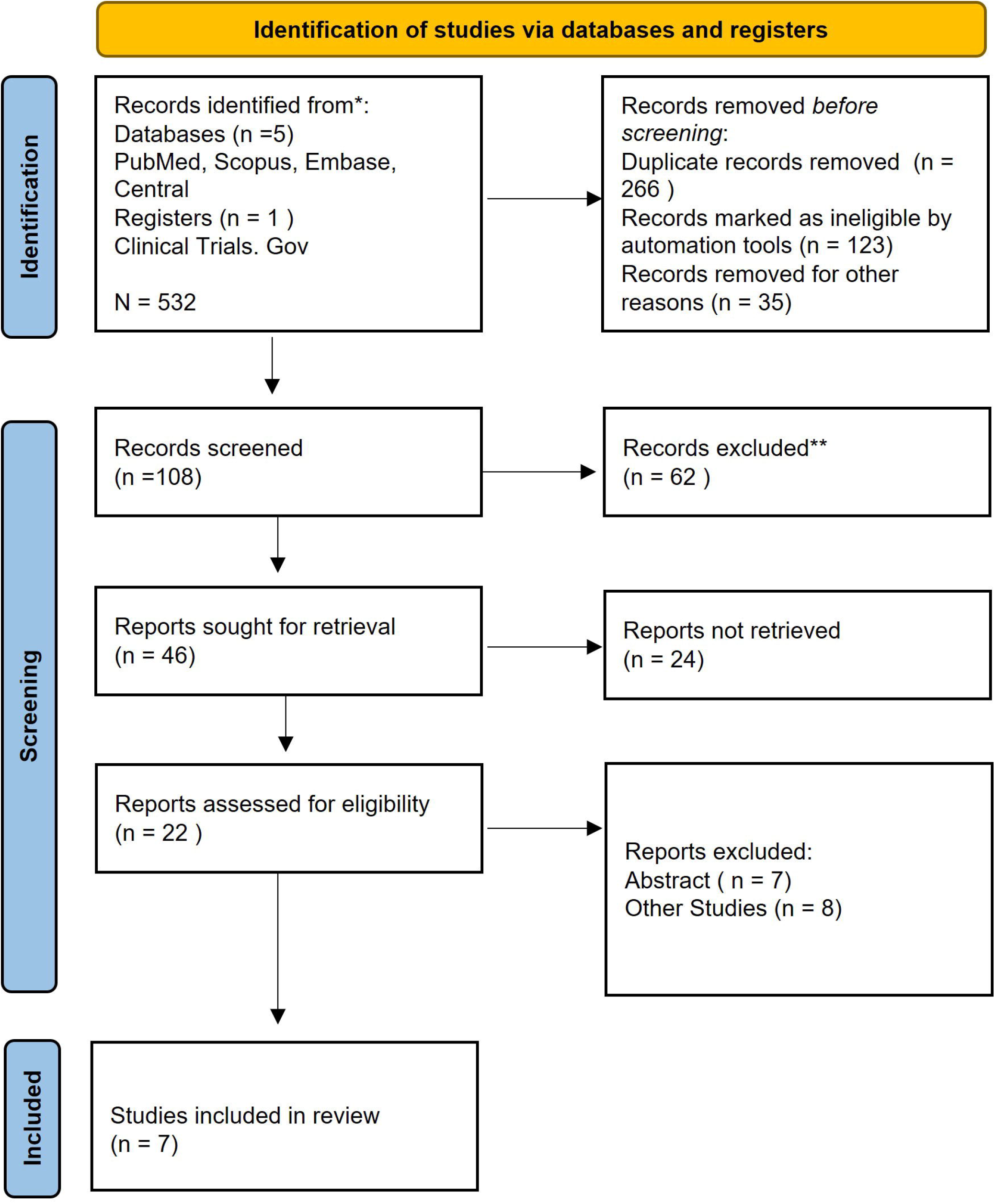
PRISMA Flow Diagram

### Dysphagia Improvement

All seven randomized controlled trials reported outcomes on dysphagia improvement following POEM versus LHM. As shown in **Figure 2**, the pooled analysis demonstrated no statistically significant difference between the two interventions. The overall log odds ratio was **0.14 (95% CI −0.32 to 0.59; *p* = 0.55)**, indicating comparable efficacy of POEM and LHM in relieving dysphagia. Individual trials varied in effect estimates, with some favoring POEM and others favoring LHM; however, the consistency across studies was supported by low-to-moderate heterogeneity (**I**^**2**^ **= 25.7%**). These findings suggest that both procedures provide substantial and durable symptomatic relief in patients with achalasia.

**Figure 2.**
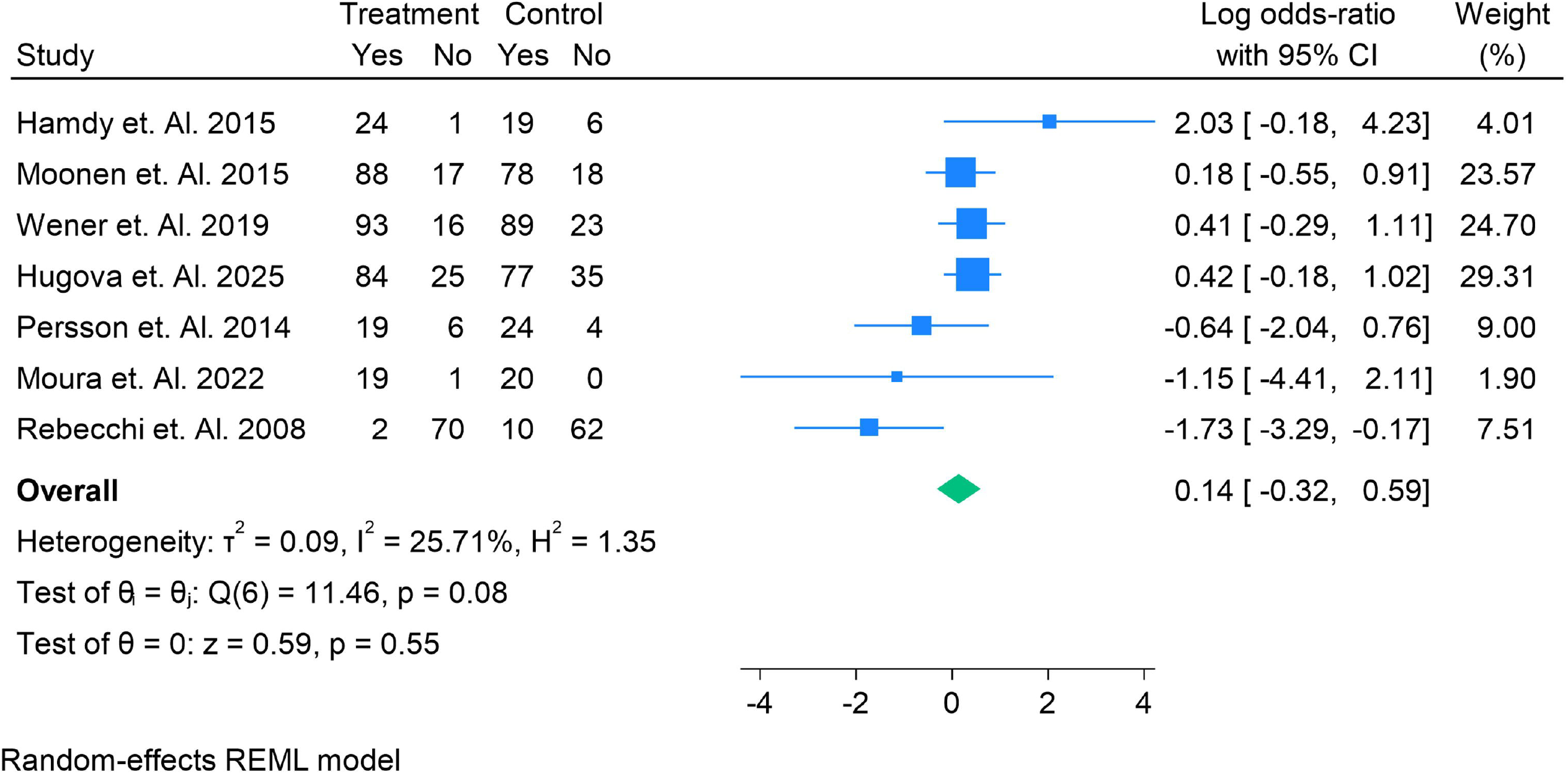
Odds ratio of the number of patients reduced in dysphagia in the LHM vs POEM

### Gastroesophageal Reflux Disease (GERD)

Five randomized controlled trials reported on postoperative GERD outcomes. As illustrated in **Figure 3**, the pooled analysis demonstrated a trend toward higher GERD incidence after POEM compared with LHM, although this did not reach statistical significance. The overall log odds ratio was **0.59 (95% CI −0.08 to 1.25; *p* = 0.08)**, suggesting a tendency favoring LHM in reducing reflux. Heterogeneity was low to moderate (**I**^**2**^ **= 29.5%**), indicating consistent findings across studies. While some trials showed a notable excess of GERD in the POEM group, others reported comparable rates between procedures.

**Figure 3.**
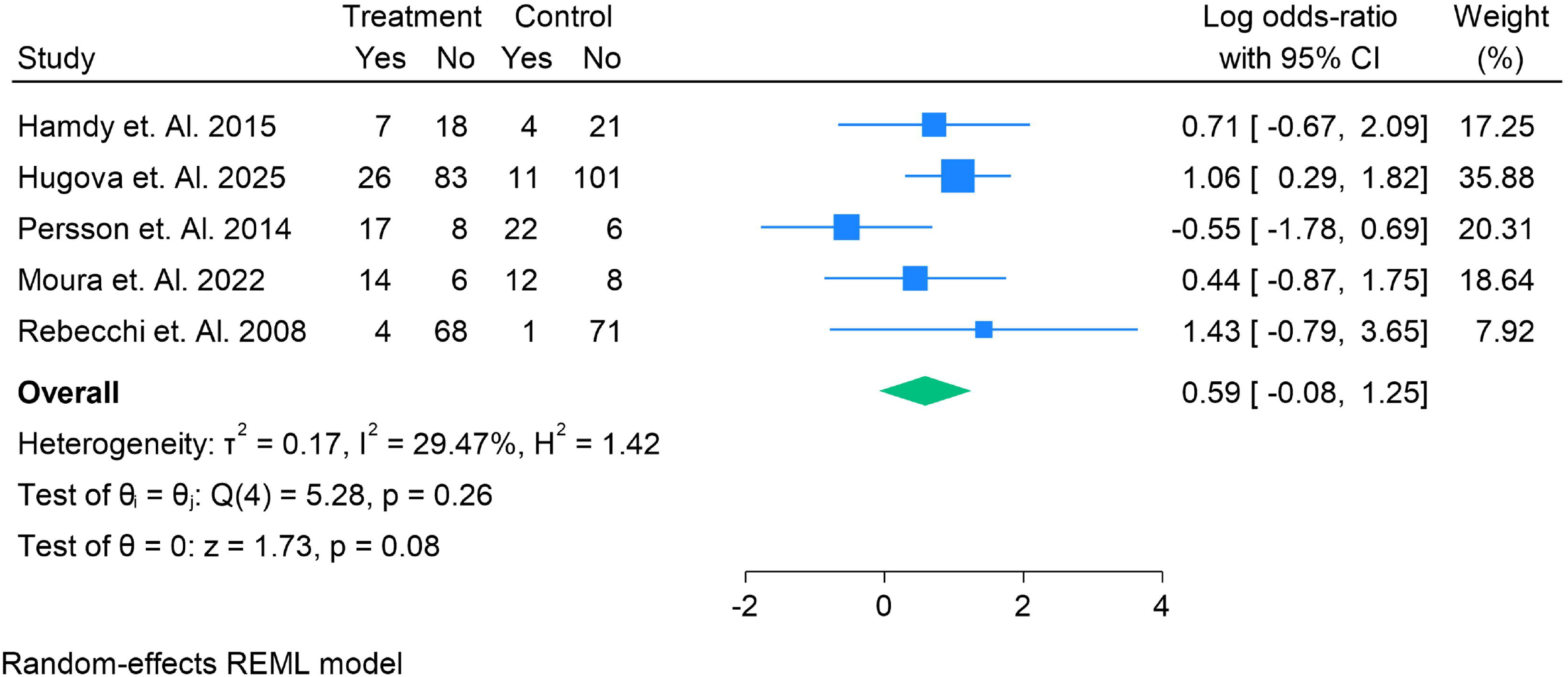
Odds ratio of the number of patients reduced in GERD in the LHM vs POEM

### Clinical Remission

Three randomized controlled trials assessed clinical remission following intervention. As shown in **Figure 4**, the pooled analysis indicated no statistically significant difference between POEM and LHM. The combined log odds ratio was **0.39 (95% CI −0.06 to 0.84; *p* = 0.09)**, suggesting a trend toward improved clinical remission with POEM, though this did not reach significance. Importantly, heterogeneity across studies was negligible (**I**^**2**^ **= 0%**), reflecting consistent results across trials. These findings indicate that both POEM and LHM achieve high and durable remission rates in patients with achalasia, with no clear superiority demonstrated.

**Figure 4.**
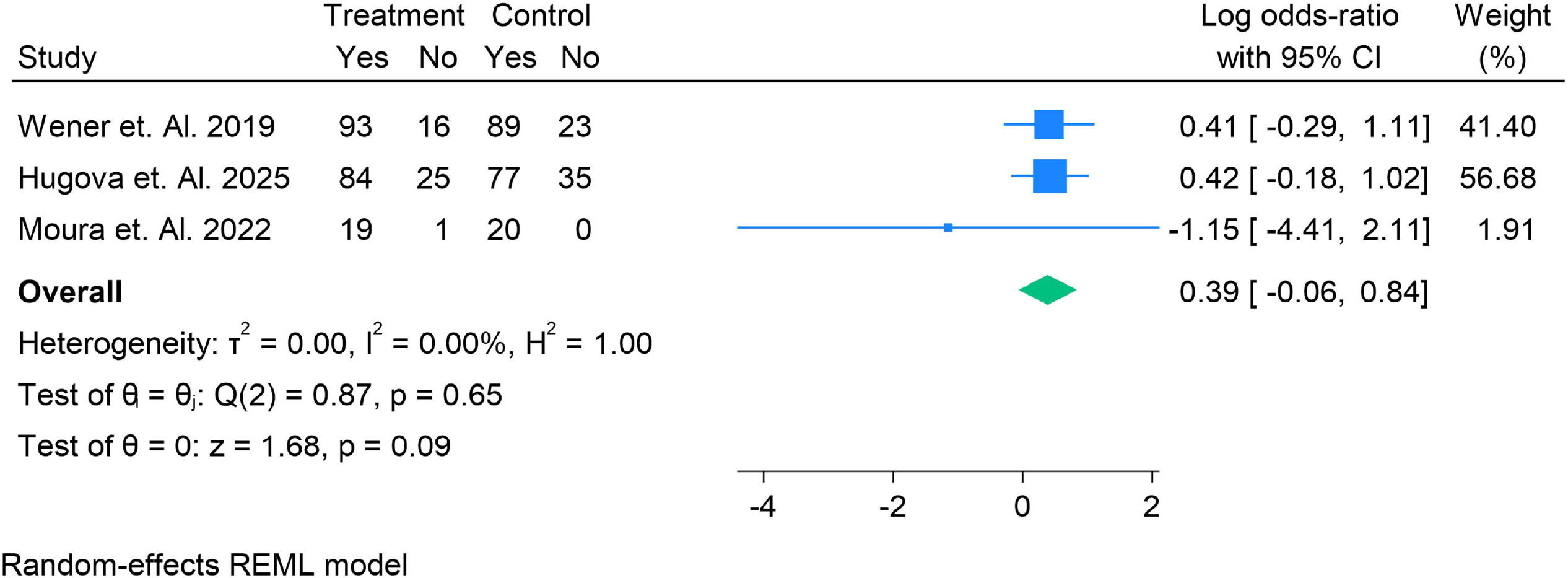
Odds ratio of the number of patients who had Clinical Remission in the LHM vs POEM

### pH Reduction (Acid Exposure)

Three randomized controlled trials reported on reduction in esophageal pH levels as an objective measure of gastroesophageal reflux. As shown in **Figure 5**, the pooled analysis demonstrated a statistically significant difference favoring LHM over POEM. The overall log odds ratio was **0.75 (95% CI 0.18 to 1.33; *p* = 0.01)**, indicating that patients undergoing POEM were more likely to develop pathological acid exposure compared with LHM. Heterogeneity was low (**I**^**2**^ **= 20.4%**), suggesting consistent results across included studies. These findings confirm that POEM, while effective for dysphagia, is associated with a higher risk of abnormal reflux on pH monitoring.

**Figure 5.**
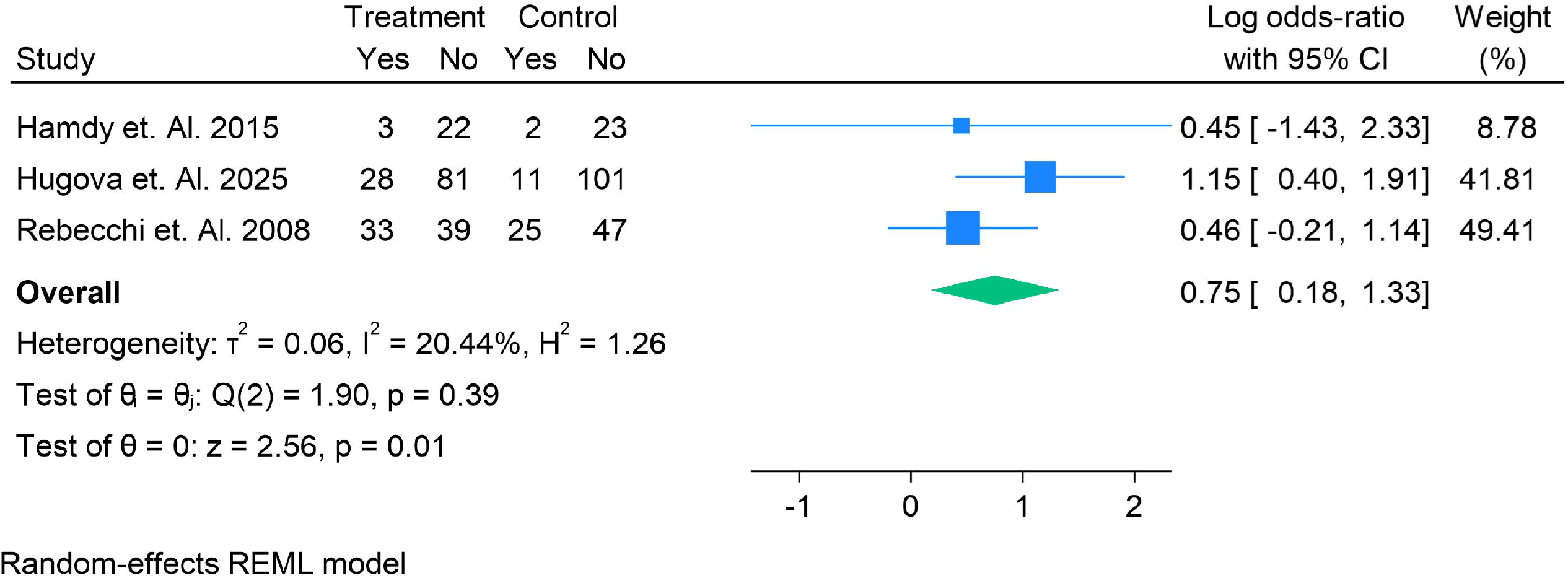
Odds ratio of the number of patients who had Reduction in pH levels in the LHM vs POEM

## Discussion

In this meta-analysis of 7 randomized controlled trials involving 900 achalasia patients (465 POEM; 465 Laparoscopic Heller Myotomy [LHM]), we found that POEM and LHM offer **comparable relief of dysphagia**; however, POEM is associated with **higher incidence of GERD symptoms**, especially when assessed objectively (pH reduction, esophagitis). Clinical remission also showed a trend favoring POEM but did not reach statistical significance; mortality remains very rare and did not differ between groups. These findings add RCT-based precision to what has been largely observational or short-term in prior literature.

When compared with **previous meta-analyses**, our results are broadly consistent. Schlottmann et al. (2018) performed a large meta□analysis including 53 LHM studies and 21 POEM studies, finding slightly better dysphagia relief with POEM at 12 and 24 months but significantly higher GERD risk (symptomatic, erosive, or via pH monitoring) after POEM [30]. Our study’s dysphagia outcome (log OR ∼0) matches the notion of **non**D**inferiority** of POEM in symptom relief, while our GERD outcomes mirror their observations of elevated reflux risk. Similarly, a more recent meta-analysis by Eltelbany et al. (2021) of long-term outcomes confirmed both modalities maintain efficacy over years, but GERD remains more frequent after POEM [31]. The systematic review by Kumar et al. (2023) also emphasized that while POEM offers minimally invasive advantages and high short-term success, its reflux risk is greater when compared to LHM (especially because LHM is routinely paired with fundoplication) [32].

Our findings refine previous knowledge by relying only on RCTs, reducing bias inherent in retrospective observational studies, and standardizing outcomes like “dysphagia improvement”, clinical remission, GERD (both subjective, symptomatic and objective), and mortality. This gives stronger internal validity, though at some cost of sample size and diversity.

### Clinical Implications

Given that dysphagia is the primary symptom driving therapy, the comparable relief between POEM and LHM suggests that POEM is a valid alternative. In patients for whom surgical access is more complicated, or in centers with strong endoscopy expertise, POEM may offer logistical advantages (shorter recovery, less external incisions). However, the elevated risk of GERD—especially when evaluated via endoscopy or pH monitoring— must be clearly communicated to patients. Long-term risks of reflux (esophagitis, Barrett’s esophagus) remain insufficiently characterized, particularly beyond 5 years, so patient selection and surveillance are important.

## Strengths and Limitations

Strengths of our meta-analysis include the exclusive use of randomized trials, balanced numbers in both arms, inclusion of objective GERD measures (pH, endoscopy), and homogeneity of clinical remission estimates (I^2^ = 0% for that outcome). Limitations include: follow-up durations in several RCTs are relatively short, particularly for GERD and mortality outcomes; sample sizes, while nontrivial, remain modest compared to meta-analyses combining observational studies; heterogeneity in definition of GERD (symptoms, endoscopy, pH) and remission criteria; possible learning curve effects for POEM in earlier studies; and many trials originate from high-volume expert centers—generalizability to broader practice may be limited.

## Comparison with Other Studies & Meta-Analyses

Other meta-analyses (e.g. by Schlottmann et al., Eltelbany et al., Kumar et al.) similarly showed that POEM and LHM are approximately equal in relieving dysphagia, with POEM possibly slightly better in short-term, but gastroesophageal reflux outcomes more problematic in POEM. Schlottmann’s predicted probabilities: ∼93.5% POEM vs ∼91.0% LHM at 12 mo, and similar at 24 mo [34]. Our dysphagia results approximate non-inferiority, though our pooled ORs did not show a statistically significant superiority of POEM. The meta-analysis by Shiu et al. (2022) and network meta-analyses also suggest that LHM + partial fundoplication reduces risk of pathologic reflux compared to POEM [33].

## Future Directions

For future research, longer-term RCTs (5+ years) are needed to assess not only symptom relief and remission but also the incidence and severity of GERD, its management, and its consequences (e.g., Barrett’s, esophageal adenocarcinoma). Standardization of outcome definitions (especially for GERD) will enhance comparability. More trials stratified by achalasia subtype (I, II, III), prior treatment status, age, and disease duration may help identify which subgroups benefit more from one procedure vs the other. Also, trials assessing cost-effectiveness, patient-reported quality of life, and impact on nutrition and long-term esophageal function are valuable.

## Conclusion

In conclusion, our RCT□based meta□analysis supports that POEM and LHM are similarly effective in relieving dysphagia and achieving clinical remission, but POEM is associated with higher risks of GERD (particularly via objective measures). For patients and clinicians, the trade-off is between less invasiveness vs higher reflux risk — informed consent and careful long-term follow-up are essential. Your work helps strengthen the evidence base by focusing on randomized data.

## Data Availability

supplementary file

## Conflict of Interest

*The authors certify that there is no conflict of interest with any financial organization regarding the material discussed in the manuscript*.

## Funding

*The authors report no involvement in the research by the sponsor that could have influenced the outcome of this work*.

## Authors’ contributions

*All authors contributed equally to the manuscript and read and approved the final version of the manuscript*.

## Notes

### Competing Interest Statement

The authors have declared no competing interest.

### Clinical Protocols

https://www.crd.york.ac.uk/PROSPERO/view/CRD420251148413

### Funding Statement

none

