## supplementary file for "Peroral Endoscopic Myotomy Versus Laparoscopic Heller Myotomy for Achalasia: A Meta-Analysis of Randomized Controlled Trials"

Supplementary Files

Search String

("Achalasia"[MeSH Terms] OR achalasia[tiab]) AND (("Myotomy"[MeSH Terms] OR myotomy[tiab] OR "Heller Myotomy"[tiab] OR "Laparoscopic Myotomy"[tiab] OR "Laparoscopic Heller Myotomy"[tiab] OR "Laparoscopic Myotomy and Fundoplication"[tiab]) OR ("Peroral Endoscopic Myotomy"[tiab] OR POEM[tiab] OR "Endoscopic Myotomy"[tiab]))

Table S1. Summary Findings of each Study

| Ref Number | Author year | Country | Total Population | Total Male | Total Female | Previous Treatment | Intervention Name | Intervention Number | Control Name | Control Number | Grade | Main Finding |
| --- | --- | --- | --- | --- | --- | --- | --- | --- | --- | --- | --- | --- |
| 14 | Hamdy et. Al. 2015 | Egypt | 50 | 13 | 37 | N/A | LMH | 25 | POEM | 25 | High | **LEM achieved higher dysphagia relief and better manometric improvement than EPD in early achalasia, with similar complication rates but higher cost; EPD was cheaper overall.** |
| 15 | Moonen et. Al. 2015 | Spain | 201 | 117 | 84 | N/A | LMH | 105 | POEM | 96 | High | At ≥5 years, **LHM and PD achieve comparable success** (≈84% vs 82%); **about 25% of PD patients require redilation**, and complication profiles differ but no clear long-term functional differences |
| 16 | Wener et. Al. 2019 | Belgium | 221 | 128 | 93 | 79 | LMH | 109 | POEM | 112 | High | POEM was noninferior to LHM+Dor for 2-year symptom control, but POEM had more endoscopic reflux esophagitis during follow-up |
| 17 | Hugova et. Al. 2025 | Belgium | 221 | 128 | 93 | 79 | LMH | 109 | POEM | 112 | High | **POEM was non-inferior to LHM + Dor fundoplication for 5-year symptom control, but had higher long-term objective reflux (pH-metry) and a numerically higher esophagitis rate.** |
| 18 | Persson et. Al. 2014 | Sweden | 53 | 23 | 30 | N/A | LMH | 25 | POEM | 28 | High | Over ≥5 years, **laparoscopic myotomy was superior to pneumatic dilatation in reducing treatment failures**, though with higher initial direct medical costs. |
| 19 | Moura et. Al. 2022 | Brazil | 40 | 26 | 14 | N/A | LMH | 20 | POEM | 20 | High | **POEM and LM-PF achieved comparable 12-month clinical success and objective improvement, but POEM had significantly higher post-procedure GER/erosive esophagitis.** |
| 20 | Rebecchi et. Al. 2008 | Italy | 114 | 75 | 69 | 19 | LMH | 72 | POEM | 72 | High | Both Heller + Dor and Heller + Nissen controlled GER long-term, but **dysphagia recurrence was significantly lower with Dor** (2.8% vs 15% at ~5 years), supporting Dor as the preferred wrap. |

Figure S1. Risk of Bias

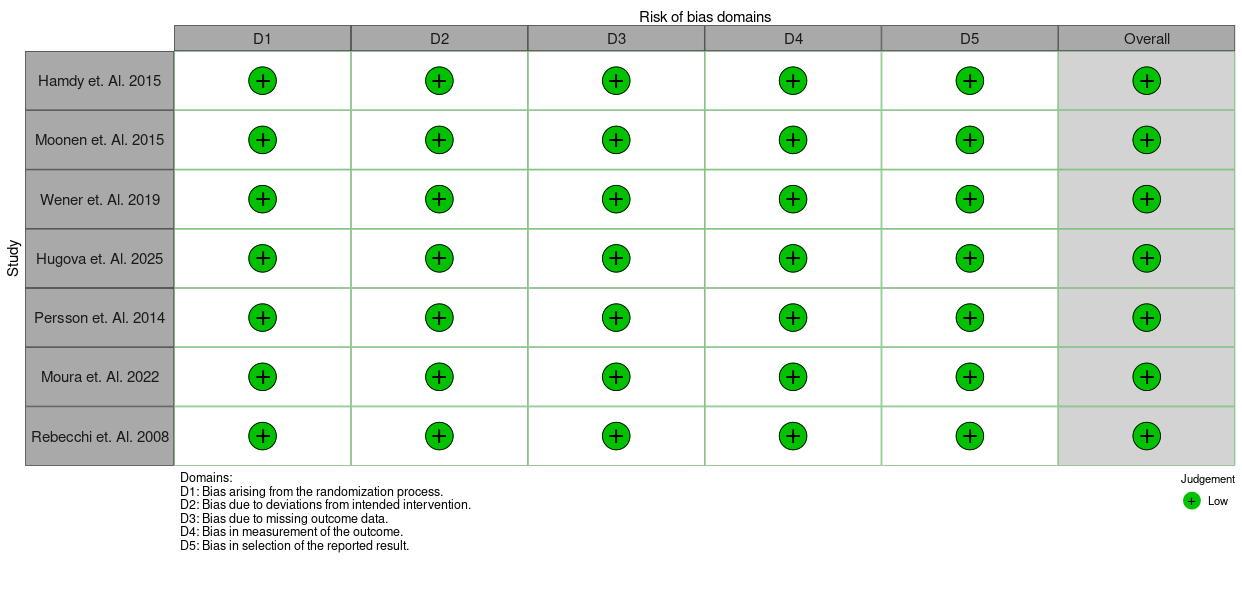
